# Maternal epigenetic age acceleration in early pregnancy, gestational duration, and spontaneous preterm birth among Black women in the NuMom2b study

**DOI:** 10.64898/2026.10.22.26366150

**Authors:** Pamela Scorza, Haotian Wu, Paolo Reho, Yihong Zhao, Veronica Barcelona

## Abstract

**Importance:** Preterm birth is a leading cause of neonatal morbidity and mortality and disproportionately affects Black birthing people in the United States. DNA methylation–based measures of biological aging may capture physiologic dysregulation relevant to birth timing. We aim to determine whether maternal epigenetic age acceleration in early pregnancy is associated with gestational duration, spontaneous preterm birth (sPTB), and small for gestational age (SGA) among Black nulliparous pregnant individuals.

**Methods:** This is a secondary data analysis of the prospective, multicenter Nulliparous Pregnancy Outcomes Study: Monitoring Mothers-to-Be (nuMoM2b). Participants enrolled in eight US clinical research sites from 2010 to 2014 were followed through delivery; DNAm arrays were processed in 2024, and the current analysis was conducted in 2026. Participants were nulliparous individuals with singleton pregnancies who self-identified as Black. Maternal peripheral blood DNA methylation was measured at 6-13 weeks’ gestation using the Illumina MethylationEPIC array. The primary aging metric was standardized GrimAge2 age acceleration. Principal Components versions of the Horvath, Hannum, PhenoAge, GrimAge, and DNAmTL clocks, and DunedinPACE, were used in additional analyses. Delivery data was abstracted from medical records in the NuMom2b study. Outcomes were gestational duration in days, based on the best obstetric estimate, adjudicated sPTB before 37 completed weeks, and SGA. Primary linear and logistic regression models adjusted for maternal age, smoking, gestational age at blood draw, and estimated blood cell composition. Secondary models added education and body mass index (BMI) as additional covariates. Fetal sex interaction effects were tested.

**Results:** Among 931 participants (mean age = 23.46 [SD=5.33] years), 55 (5.9%) experienced sPTB. Each 1-SD higher GrimAge2 acceleration was associated with 1.32 fewer days of gestation (β, −1.32 days; 95% CI, −2.36 to −0.28; *P*=.01) and higher odds of sPTB (odds ratio, 1.51; 95% CI, 1.13-2.02; *P*=.01). Results were slightly attenuated but remained significant for sPTB when controlling for education and BMI. Epigenetic age acceleration was not associated with SGA. Results were not significantly different by fetal sex.

**Conclusions and Relevance:** Greater first-trimester maternal GrimAge2 acceleration was associated with shorter gestation and 51% higher odds of sPTB but was not associated with SGA among Black nulliparous participants. The association with sPTB persisted after adjustment for socioeconomic and metabolic characteristics, warranting further evaluation of whether GrimAge2 improves early-pregnancy prediction of sPTB value beyond these factors.

## Background

Preterm birth (PTB), defined as delivery before 37 completed weeks of gestation, remains a leading cause of neonatal morbidity and mortality worldwide and is a major contributor to long-term neurodevelopmental and cardiometabolic risk across the life course.^1,2^ In the United States, the burden of PTB is inequitably distributed, with rates consistently higher among Black birthing people than among other racial or ethnic groups.^3^ Rather than innate biological differences, these disparities are understood to reflect the cumulative effects of structural, social, and environmental inequities^4^ that may become biologically embedded in ways relevant to birth timing. Biological indicators associated with PTB could ultimately improve risk assessment and provide tools for evaluating interventions and policies intended to improve birth outcomes.

DNA methylation (DNAm)-based epigenetic clocks derive measures of biological aging from methylation patterns across the genome.^5^ Epigenetic age acceleration— the extent to which estimated age exceeds chronological age— has been associated with morbidity, mortality, and physiological dysregulation in non-pregnant populations.^6^ During pregnancy, maternal epigenetic aging may reflect cumulative stress exposures, inflammation, and metabolic dysfunction,^7–10^ processes potentially relevant to shortened gestation and spontaneous PTB (sPTB).

Evidence linking maternal epigenetic age acceleration to birth outcomes is growing but mixed. In the largest study to date, conducted in the Norwegian Mother, Father and Child Cohort Study (MoBa; n=2,198), higher second-trimester maternal epigenetic age acceleration in five of six epigenetic clocks, was associated with shorter gestation and a higher risk of sPTB.^11^ Smaller studies have reported associations between GrimAge acceleration and PTB in a high-risk clinical sample (n=163),^12^ between GrimAge acceleration and shorter gestation in the Healthy Babies Before Birth cohort (n=77),^13^ and between GrimAge2 acceleration and a composite indicator of pregnancy complications, which included preterm birth (n=45).^14^ An earlier study associated first-trimester Horvath epigenetic age acceleration with PTB, with the association primarily driven by participants who self-identified as African American.^15^ In contrast, a study in the Philippines (n=296) found little evidence of association across 15 clocks and multiple birth outcomes.^16^ Collectively, these findings suggest that maternal epigenetic aging may be relevant to gestational duration and sPTB, while underscoring the need for replication in larger, well-characterized and diverse populations.

Despite growing interest in this area, large-scale studies remain scarce, and none have focused specifically on Black pregnant individuals. This gap matters given the disproportionate burden of PTB among Black birthing populations in the United States and the potential for structurally patterned exposures to shape biological aging processes before and during pregnancy. The Nulliparous Pregnancy Outcomes Study: Monitoring Mothers-to-Be (NuMoM2b) provides a unique opportunity to address this gap. Genome-wide DNAm profiles were generated from stored first-trimester blood samples for more than 900 Black participants using the Illumina EPIC v2 array and linked to rigorously characterized pregnancy outcomes, including ultrasound-dated gestational age and adjudicated sPTB.^17^

We examined whether maternal epigenetic age in early pregnancy was associated with gestational duration and sPTB among Black participants in NuMoM2b. We also tested whether maternal epigenetic age acceleration in early pregnancy was associated with small for gestational age (SGA) and whether associations differed by fetal sex, given evidence for sex-specific developmental and epigenetic processes beginning early in gestation.^18–20^

## Methods

### Study design and population

We conducted a secondary analysis within the Nulliparous Pregnancy Outcomes Study: Monitoring Mothers-to-Be (NuMoM2b), a prospective multicenter cohort study of nulliparous pregnant individuals from eight academic medical centers in the United States between 2010 and 2014. Verbal or written informed consent was obtained from participants during recruitment. Whole blood of nulliparous pregnant individuals was collected during early pregnancy between 6+0 and 13+6 weeks of gestation. Behavioral, demographic, and clinical data were collected, and electronic health records data were utilized for birth outcomes. Additional details are provided in the nuMoM2b study protocol.^17^ Participants with multiple gestations, medically indicated terminations, medically indicated PTB, extreme implausible gestational ages at delivery (<22 or >43 weeks), or samples failing DNAm quality control were excluded. The analytic sample included 931 self-identified Black participants with available DNAm data from maternal peripheral blood collected during pregnancy and complete delivery outcome data and covariate data. Study procedures for the current analyses were approved by the Columbia University Institutional Review Board. (#AAAU0215)

### DNA methylation profiling and epigenetic clock estimation

Genome-wide DNAm profiling was performed using the Illumina MethylationEPIC v2 array. DNAm data were preprocessed using the MethylCallR R package.^21^ Additional preprocessing details are provided in Zhao et al 2026.^22^ Raw IDAT files from 1,073 samples were imported for quality assessment. Samples were excluded if their median log_2_ signal intensity was below 10.5 or if more than 1% of probes had detection P values greater than 0.01. Probes were excluded based on detection performance, bead count, genomic location, sequence characteristics, reproducibility, and alignment quality. Specifically, we removed probes with detection P values greater than 0.01 in more than 1% of samples, bead counts below 3 in more than 5% of samples, non-CG targets, chromosome Y locations, common SNPs in African-ancestry populations according to the SeSaMe AFR manifest, evidence of cross-hybridization based on BLAT, low reproducibility in the EPIC v2 manifest, or unsuccessful or discordant mapping to GRCh38 relative to the SeSaMe manifest. Duplicated EPIC v2 probes were additionally removed using MethylCallR reference data to facilitate cross-platform compatibility. These procedures excluded 3 samples and 102,769 probes, leaving 834,231 high-confidence CpG sites. Data were normalized using normal-exponential out-of-band correction with an offset of 100 and without interpolation of sex-chromosome probes. Technical variation associated with known batch factors, including Sentrix barcode and sample section, was addressed using MeCall.RemoveBatch(), with PTB status protected during batch correction. We excluded 11 outliers with atypical methylation profile based on Mahalanobis-distance.

We selected GrimAge2 acceleration as our primary epigenetic measure because it is an updated version of GrimAge and incorporates DNAm-based estimates of inflammatory and metabolic markers, including C-reactive protein and hemoglobin A1c, that may be relevant to birth timing. It has shown stronger associations with mortality and age-related morbidity than the original GrimAge.^7^ DunedinPACE as well as principal component (PC)-based versions of additional established clocks were also analyzed for comparison with other studies, including Horvath, Hannum, PhenoAge, GrimAge, and DNAmTL. PC-based clocks were selected to improve technical reliability and reduce noise relative to traditional single-estimate approaches.^23^ GrimAge2 was calculated using the *EpigeneticAgePipeline* package,^24^ while PC epigenetic clock measures and DunedinPACE were calculated using the *methylCIPHER* package.^25^ For each clock, epigenetic age acceleration was defined as the residual from a regression of epigenetic age on chronological age. Resulting residuals were standardized to a mean of 0 and standard deviation (SD) of 1, such that estimates represent associations per 1-SD higher age acceleration.

### Outcomes

The primary outcomes were 1) Gestational duration: gestational age at delivery in days, based on the best obstetric estimate. 2) sPTB: spontaneous labor and delivery before 37 completed weeks of gestation. In NuMom2b, PTB was categorized as spontaneous or indicated. sPTB was defined as delivery of a liveborn or stillborn infant from 20 weeks 0 days through 36 weeks 6 days of gestation following spontaneous preterm labor or premature rupture of membranes.

Preterm labor was defined by either (1) spontaneous uterine contractions preceding rupture of membranes and resulting in delivery or (2) among participants hospitalized for contractions, an increase in cervical dilation of at least 1 cm or an increase in effacement, cervical dilation greater than 2 cm, or cervical effacement greater than 80%. Secondary analyses also examined small for gestational age (<10^th^ percentile), calculated using Oken criteria.^26,27^

### Covariates

Primary models adjusted for maternal chronological age at enrollment, gestational age at blood draw, smoking during pregnancy (any versus none), fetal sex, and cell type composition. Pre-pregnancy body mass index and educational attainment were considered in sensitivity analyses.

### Statistical analysis

For each clock, associations with gestational duration were estimated using multivariable linear regression. Regression coefficients and 95% confidence intervals (CIs) represent the mean difference in gestational duration (days) per 1-SD higher age acceleration.

Associations with sPTB were estimated using multivariable logistic regression. Odds ratios (ORs) and 95% CIs represent the relative odds of PTB per 1-SD higher age acceleration. GrimAge2 was the prespecified primary clock; its results are presented with nominal p-values. The other clocks were considered for comparison with other studies, and their p-values were adjusted for multiple testing using the false discovery rate (FDR) method, with a threshold of .05.

### Effect modification by fetal sex

To evaluate potential heterogeneity by fetal sex, we fit interaction models including a product term between age acceleration and fetal sex. Sex-specific estimates and interaction p-values were reported.

## RESULTS

The analytic sample included 931 Black nulliparous participants (mean age 23.46 years). Mean gestational duration was 274.38 days. Fifty-five births (5.9%) were preterm, and 133 infants (14.3%) were small for gestational age (Table 1). Infants included 460 females (49.4%) and 471 males (50.6%).

**Table 1.** Participant and birth characteristics (N = 931)

| Characteristic | Value |
| --- | --- |
| Maternal age, mean (SD), years | 23.46 (5.33) |
| BMI, mean (SD) | 28.86 (7.90) |
| Smoking status, n (%) |  |
| No | 832 (89.4) |
| Yes | 99 (10.6) |
| Education, n (%) |  |
| <High school | 155 (16.6) |
| High school or GED | 242 (26.0) |
| Some college | 277 (29.8) |
| Associate/tech degree | 89 (9.6) |
| College degree | 103 (11.1) |
| Post-graduate degree | 65 (7.0) |
| Fetal sex, n (%) |  |
| Female | 460 (49.4) |
| Male | 471 (50.6) |
| Gestational duration, mean (SD), days | 274.38<br>(14.32) |
| Preterm birth, n (%) | 55 (5.9) |
| Small for gestational age, n (%) | 133 (14.3) |

The epigenetic age-estimating clocks were positively correlated with chronological age (r = 0.712–0.798), whereas DNAmTL was inversely correlated with chronological (r = −0.583); all had *P* < .001 (Figure 1).

**Figure 1.**
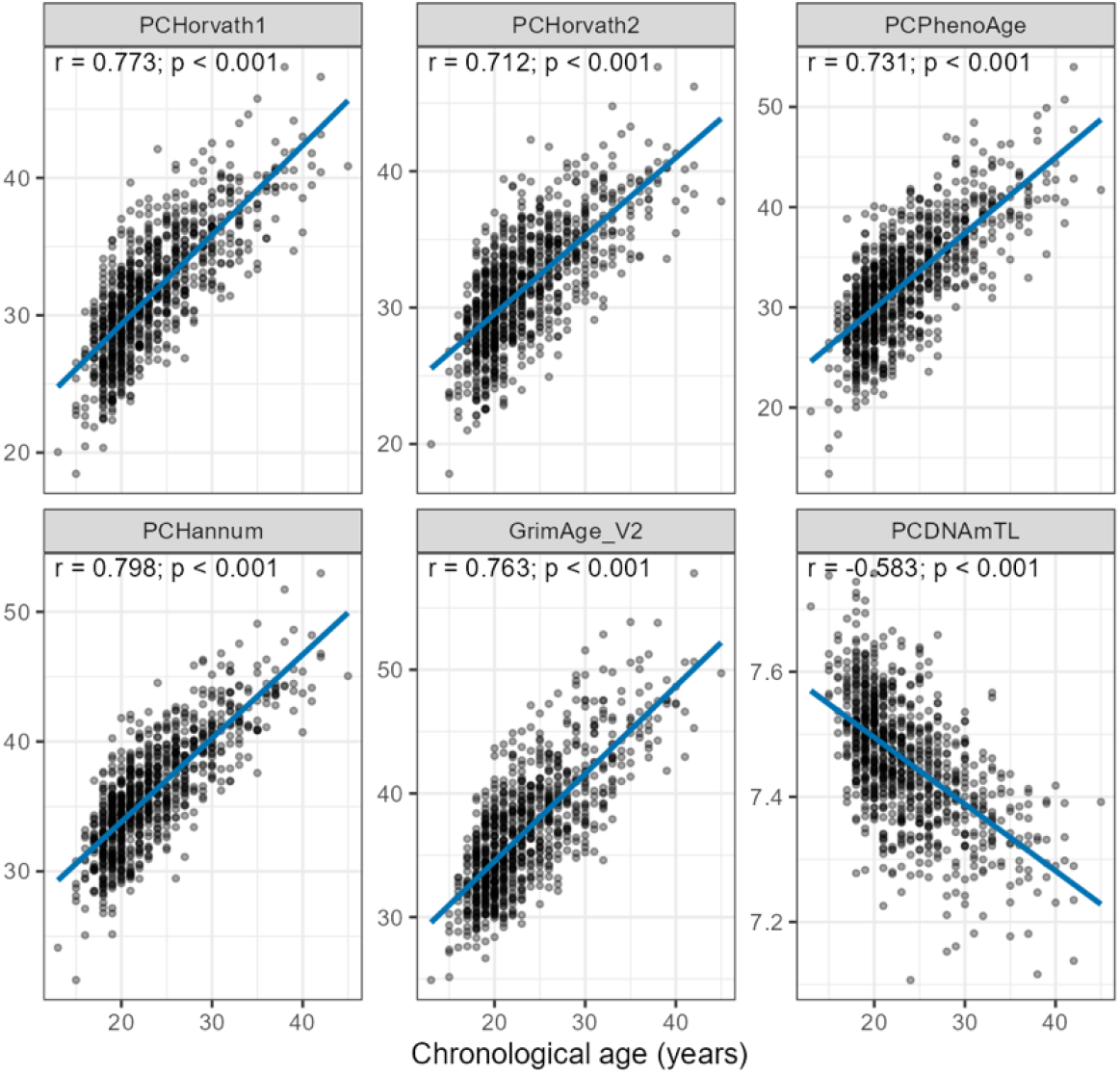
Associations of DNA methylation-based clock estimates with chronological age

In models adjusted for maternal age, gestational age at blood draw, fetal sex, smoking, and estimated cell composition, greater PCGrimAge2 acceleration was associated with shorter gestational duration and higher odds of spontaneous preterm birth (Figure 2). Each SD increase in PCGrimAge2 acceleration was associated with a 1.32-day reduction in gestational duration (95% CI, -2.36 to -.28 days; P = .01) and 51% higher odds of spontaneous preterm birth (Adjusted Odds Ratio [aOR] 1.51; 95% CI, 1.13–2.02; P = .01). PCGrimAge2 acceleration was not associated with small-for-gestational-age birth (OR, 1.07; 95% CI, .87–1.32; P = .53).

**Figure 2:**
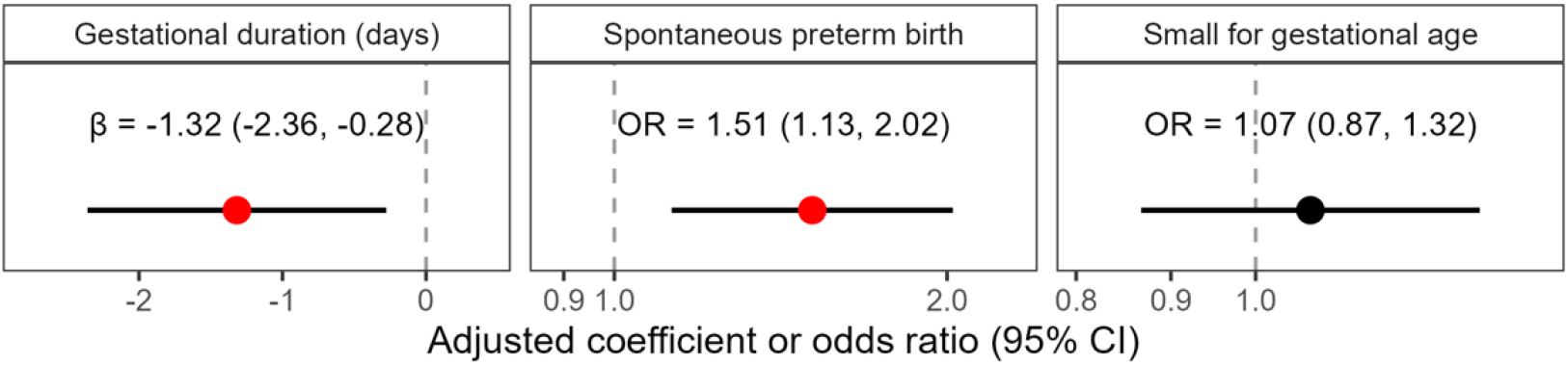
GrimAge2 Acceleration and Perinatal Outcomes, adjusted for maternal age, fetal sex, smoking, gestational age at blood draw, and cell type composition.

After further adjustment for education and BMI, the effects were slightly attenuated, and the p-value of the association with gestational duration reached p = 0.05 (Table 2).

**Table 2.** GrimAge2 Acceleration and Perinatal Outcomes, additionally adjusted for education and BMI.

|  | Difference (95% CI) | p-value |
| --- | --- | --- |
| Gestational duration | -1.10 (-2.19, 0.00) | 0.05 |
|  | OR (95% CI) | p-value |
| Small for gestational age | 1.09 (0.87, 1.36) | 0.45 |
| Preterm birth | 1.40 (1.03, 1.91) | 0.03 |

There was no evidence that associations of PCGrimAge2 acceleration with birth outcomes differed by fetal sex (likelihood ratio test for interaction p = 0.96 for small-for-gestational-age birth, p = 0.99 for gestational duration, and p = 0.21 for spontaneous preterm birth). Among female infants, greater PCGrimAge2 acceleration was associated with shorter gestational duration (β = −1.74 days; 95% CI, −3.18 to −0.31 days; P = .02) and higher odds of spontaneous preterm birth (aOR, 1.75; 95% CI, 1.18–2.59; P = .01). Corresponding estimates among male infants were −0.88 days (95% CI, −2.39 to 0.63 days; P = .25) and 1.21 (95% CI, 0.78–1.88; P = .39), respectively. Associations with small-for-gestational-age birth were near the null among both female and male infants (Supplementary Table 1).

Among the remaining DNA methylation clocks, greater age-adjusted PCDNAmTL was associated with lower odds of spontaneous preterm birth (aOR, 0.68; 95% CI, 0.51–0.89; P = .01; FDR-adjusted p= .03). PCDNAmTL was not associated with gestational duration (β = 0.35 days; 95% CI, −0.60 to 1.31 days) or small-for-gestational-age birth (aOR, 0.95; 95% CI, 0.78– 1.15). Neither age acceleration measured using PCHorvath1, PCHorvath2, PCPhenoAge, or PCHannum nor DunedinPACE was associated with any of the 3 birth outcomes; all corresponding 95% CIs included the null (Supplementary Table 2).

## Discussion

In this cohort of over 900 Black nulliparous participants, each standard deviation increase in first-trimester maternal PCGrimAge2 acceleration was associated with shorter gestation and approximately 51% higher odds of sPTB. The association with sPTB persisted after adjustment for BMI and education, suggesting that GrimAge2 captures variation in sPTB risk beyond these measured characteristics. We found no association with SGA and no evidence that associations differed by fetal sex. These findings extend evidence linking maternal epigenetic aging with birth timing to a population that bears a disproportionate burden of preterm birth but has not been the focus of prior large studies of epigenetic clocks in pregnancy.

Our findings are broadly consistent with those of Magnus et al., who reported an association between maternal PCGrimAge acceleration and sPTB in the Norwegian Mother, Father and Child Cohort Study (OR per SD, 1.36; 95% CI, 1.09–1.70). In our sample, we focused our primary analysis on the newer PCGrimAge2 and found that the association with sPTB and found the correlation to be similar (OR per SD, 1.40; 95% CI 1.03-1.91) after adjusting for education and BMI. The consistency of these estimates supports further investigation of GrimAge-based measures across pregnancy populations.

Among the additional aging measures we examined, only the DNAm-based indicator of telomere length showed a statistically significant association. This finding aligns with preliminary evidence linking shorter measured maternal leukocyte telomeres to shorter gestation among Black women^28^ and has broader context in Geronimus and colleagues’ landmark work on telomere length and weathering among midlife Black women.^29^ However, the DNAm-based indicator of telomere length is a proxy for directly measured telomere length. The concentration of our findings in the GrimAge measure and DNAmTL, with little evidence of association for PhenoAge or DunedinPACE, suggests that the observed pattern may reflect differences in the physiological information captured by these measures rather than a general distinction between chronological-age and health-related clocks. GrimAge incorporates DNAm surrogates for smoking exposure and circulating proteins related to vascular, metabolic, and stress-related physiology. Consistent with this interpretation, an earlier pregnancy study reported associations of shorter gestation with GrimAge acceleration and DNAm surrogates for plasminogen activator inhibitor-1, adrenomedullin, and cystatin C, but not with the other aging indices examined.^30^ GrimAge2 additionally incorporates DNAm estimators of C-reactive protein and hemoglobin A1c, making inflammatory and metabolic processes plausible contributors to its association with birth outcomes. These features suggest that our findings may reflect physiological variation relevant to pregnancy maintenance rather than a uniform acceleration across all dimensions of biological aging. However, the composite nature of these measures precludes identification of the processes responsible for the associations. Analyses of individual GrimAge components, direct measurements of telomere length, and repeated measurements across pregnancy would help distinguish these explanations and clarify which aspects of maternal physiology these associations represent.

The persistence of the PCGrimAge2 association after adjustment suggests that it captures variation associated with sPTB beyond that accounted for by measured BMI and education. Longitudinal analyses explicitly examining upstream exposures, DNAm, and birth outcomes will be needed to distinguish the possibilities of mediation, improved clinical prediction and potential causality.

The absence of an association with SGA suggests that the observed pattern may be more closely related to birth timing than fetal growth. We also found no evidence of interaction by fetal sex. Given the limited number of sPTB events, these analyses may have had limited power to detect differences by sex.

Strengths of this study include first-trimester blood collection, standardized DNAm measurement, and adjudicated sPTB outcomes in a well-characterized multicenter cohort. Its focus on Black participants addresses a gap in a literature drawn largely from other populations. Several limitations warrant consideration. The observational design does not establish a causal role for epigenetic aging in birth timing. Pregnancy-related physiological changes may also affect clock estimates. Although the association with sPTB persisted after adjustment for BMI and education, this analysis did not test whether GrimAge2 improves prediction beyond an early-pregnancy clinical model. That question will require explicit model comparison and validation in an independent cohort.

In summary, first-trimester maternal GrimAge2 acceleration was associated with shorter gestation and greater odds of sPTB among Black nulliparous participants, with the sPTB association persisting after adjustment for BMI and education. These results support further investigation of the processes captured by GrimAge-related measures during pregnancy and their potential value for assessing birth timing.

## Data Availability

NuMoM2b phenotype data are publicly available through the Eunice Kennedy Shriver National Institute of Child Health and Human Development Data and Specimen Hub NICHD (DASH). Epigenomic data are publicly available through the NIH Database of Genotypes and Phenotypes (dbGaP) repository under accession number phs003992.v1.p1. Both are controlled access repositories.

## Supplementary material

**Table S1.** Sex Stratified Results - GrimAge2.

| Outcome | Sex | N | Measure | Estimate (95% CI) | P-value | P-interaction* |
| --- | --- | --- | --- | --- | --- | --- |
| Small for gestational age | Female | 460 | Odds ratio | 1.12 (0.85, 1.47) | 0.42 | 0.96 |
| Small for gestational age | Male | 471 | Odds ratio | 1.01 (0.72, 1.43) | 0.94 |  |
| Gestational age at birth | Female | 460 | Beta (days) | -1.74 (-3.18, -0.31) | 0.02 | 1.00 |
| Gestational age at birth | Male | 471 | Beta (days) | -0.88 (-2.39, 0.63) | 0.25 |  |
| Spontaneous preterm birth | Female | 460 | Odds ratio | 1.75 (1.18, 2.59) | 0.01 | 0.21 |
| Spontaneous preterm birth | Male | 471 | Odds ratio | 1.21 (0.78, 1.88) | 0.39 |  |
| *Calculated from likelihood ratio test<br>Adjusted for age, infant sex, smoking, gestation age at blood draw, and cell composition |  |  |  |  |  |  |

**Table S2.** Additional Epigenetic Clocks and Perinatal Outcomes, adjusted for maternal age, infant sex, smoking, gestational age at blood draw, and cell type composition.

| Outcome | Clock | Measure | Estimate (95% CI) | P | FDR |
| --- | --- | --- | --- | --- | --- |
| Small for gestational age | PCHorvath1_AgeAccel | Odds ratio | 0.94 (0.78, 1.14) | 0.53 | 0.61 |
| Small for gestational age | PCHorvath2_AgeAccel | Odds ratio | 0.89 (0.73, 1.09) | 0.26 | 0.54 |
| Small for gestational age | PCPhenoAge_AgeAccel | Odds ratio | 0.94 (0.75, 1.18) | 0.61 | 0.61 |
| Small for gestational age | PCHannum_AgeAccel | Odds ratio | 0.89 (0.73, 1.09) | 0.26 | 0.54 |
| Small for gestational age | PCDNAmTL_AgeAccel | Odds ratio | 0.95 (0.78, 1.15) | 0.57 | 0.61 |
| Small for gestational age | dunedin_z | Odds ratio | 1.12 (0.92, 1.36) | 0.27 | 0.54 |
| Gestational age at birth | PCHorvath1_AgeAccel | Beta (days) | -0.47 (-1.40, 0.47) | 0.33 | 0.77 |
| Gestational age at birth | PCHorvath2_AgeAccel | Beta (days) | -0.17 (-1.15, 0.81) | 0.73 | 0.80 |
| Gestational age at birth | PCPhenoAge_AgeAccel | Beta (days) | -0.14 (-1.24, 0.96) | 0.80 | 0.80 |
| Gestational age at birth | PCHannum_AgeAccel | Beta (days) | -0.33 (-1.32, 0.66) | 0.51 | 0.77 |
| Gestational age at birth | PCDNAmTL_AgeAccel | Beta (days) | 0.35 (-0.60, 1.31) | 0.47 | 0.77 |
| Gestational age at birth | dunedin_z | Beta (days) | -0.77 (-1.74, 0.20) | 0.12 | 0.72 |
| Spontaneous preterm birth | PCHorvath1_AgeAccel | Odds ratio | 1.14 (0.87, 1.51) | 0.35 | 0.41 |
| Spontaneous preterm birth | PCHorvath2_AgeAccel | Odds ratio | 1.13 (0.84, 1.51) | 0.43 | 0.43 |
| Spontaneous preterm birth | PCPhenoAge_AgeAccel | Odds ratio | 1.21 (0.87, 1.67) | 0.26 | 0.39 |
| Spontaneous preterm birth | PCHannum_AgeAccel | Odds ratio | 1.21 (0.90, 1.62) | 0.21 | 0.39 |
| Spontaneous preterm birth | PCDNAmTL_AgeAccel | Odds ratio | 0.68 (0.51, 0.89) | 0.01 | 0.03 |
| Spontaneous preterm birth | dunedin_z | Odds ratio | 1.19 (0.90, 1.58) | 0.23 | 0.39 |

Adjusted for age, infant sex, smoking, gestation age at blood draw, and cell composition

## References

1. Mitha A, Chen R, Razaz N, et al. Neurological development in children born moderately or late preterm: national cohort study. BMJ. 2024;384:e075630. doi:10.1136/bmj-2023-075630

2. Ohuma EO, Moller AB, Bradley E, et al. National, regional, and global estimates of preterm birth in 2020, with trends from 2010: a systematic analysis. The Lancet. 2023;402(10409):1261–1271. doi:10.1016/S0140-6736(23)00878-4

3. Osterman MJK, Hamilton BE, Martin JA, Driscoll AK, Valenzuela CP. Births: Final Data for 2024. June 9, 2026. doi:10.15620/cdc/252440

4. Braveman P, Dominguez TP, Burke W, et al. Explaining the Black-White Disparity in Preterm Birth: A Consensus Statement From a Multi-Disciplinary Scientific Work Group Convened by the March of Dimes. Front Reprod Health. 2021;3. doi:10.3389/frph.2021.684207

5. Raj K, Horvath S. Current perspectives on the cellular and molecular features of epigenetic ageing. Exp Biol Med. 2020;245(17):1532–1542.

6. Horvath S, Raj K. DNA methylation-based biomarkers and the epigenetic clock theory of ageing. Nat Rev Genet. 2018;19(6):371–384.

7. Lu AT, Binder AM, Zhang J, et al. DNA methylation GrimAge version 2. Aging. 2022;14(23):9484–9549. doi:10.18632/aging.204434

8. Belsky DW, Caspi A, Corcoran DL, et al. DunedinPACE, a DNA methylation biomarker of the pace of aging. Elife. 2022;11:e73420.

9. Levine ME, Lu AT, Quach A, et al. An epigenetic biomarker of aging for lifespan and healthspan. Aging. 2018;10(4):573.

10. Kusters CDJ, Horvath S. Quantification of Epigenetic Aging in Public Health. Annu Rev Public Health. 2025;46(1):91–110. doi:10.1146/annurev-publhealth-060222-015657

11. Magnus MC, Lee Y, Carlsen EØ, et al. Parental epigenetic age acceleration and risk of adverse birth outcomes: the Norwegian mother, father and child cohort study. BMC Med. 2024;22(1):554. doi:10.1186/s12916-024-03780-7

12. Gascoigne EL, Roell KR, Eaves LA, Fry RC, Manuck TA. Accelerated epigenetic clock aging in maternal peripheral blood and preterm birth. Am J Obstet Gynecol. 2024;230(5):559.e1-559.e9. doi:10.1016/j.ajog.2023.09.003

13. Ross KM, Carroll JE, Horvath S, Hobel CJ, Coussons-Read ME, Dunkel Schetter C. Epigenetic age and pregnancy outcomes: GrimAge acceleration is associated with shorter gestational length and lower birthweight. Clin Epigenetics. 2020;12:1–11.

14. Panelli DM, Gladish N, Perlman NC, et al. Longitudinal Epigenetic Aging in Pregnancy and Associations with Adverse Outcomes. Obstet Gynecol. 2025;146(6):783–792. doi:10.1097/AOG.0000000000006000

15. Lancaster EE, Lapato DM, Jackson-Cook C, Strauss JF, Roberson-Nay R, York TP. Maternal biological age assessed in early pregnancy is associated with gestational age at birth. Sci Rep. 2021;11(1):15440. doi:10.1038/s41598-021-94281-7

16. Ryan CP, Rege RJ, Lee NR, et al. Maternal epigenetic clocks measured during pregnancy do not predict gestational age at delivery or offspring birth outcomes: a replication study in metropolitan Cebu, Philippines. Clin Epigenetics. 2022;14(1):78. doi:10.1186/s13148-022-01296-6

17. Haas DM, Parker CB, Wing DA, et al. A description of the methods of the Nulliparous Pregnancy Outcomes Study: monitoring mothers-to-be (nuMoM2b). Am J Obstet Gynecol. 2015;212(4):539.e1-539.e24. doi:10.1016/j.ajog.2015.01.019

18. Czamara D, Dieckmann L, Lahti-Pulkkinen M, et al. Sex differences in DNA methylation across gestation: a large scale, cross-cohort, multi-tissue analysis. Cell Mol Life Sci. 2024;81(1):177. doi:10.1007/s00018-024-05208-0

19. Bale TL. Sex differences in prenatal epigenetic programing of stress pathways. Stress. 2011;14(4):348–356.

20. Gonzalez TL, Willson BE, Wang ET, et al. Sexually dimorphic DNA methylation and gene expression patterns in human first trimester placenta. Biol Sex Differ. 2024;15(1):63. doi:10.1186/s13293-024-00629-9

21. Yang HH, Han MR. MethylCallRL: a comprehensive analysis framework for Illumina Methylation Beadchip. Sci Rep. 2024;14(1):27026. doi:10.1038/s41598-024-77914-5

22. Zhao T, Zhao Y, Reho P, et al. Epigenome-wide DNA methylation and spontaneous preterm birth among pregnant black women. Clin Epigenetics. 2026;18(1):146. doi:10.1186/s13148-026-02151-8

23. Higgins-Chen AT, Thrush KL, Wang Y, et al. A computational solution for bolstering reliability of epigenetic clocks: Implications for clinical trials and longitudinal tracking. Nat Aging. 2022;2(7):644–661.

24. Castellani DC. CastellaniLab/EpigeneticAgePipeline. Published online September 10, 2026. Accessed October 21, 2026. https://github.com/CastellaniLab/EpigeneticAgePipeline

25. Thrush KL, Higgins-Chen AT, Liu Z, Levine ME. R methylCIPHER: A Methylation Clock Investigational Package for Hypothesis-Driven Evaluation & Research. bioRxiv. Preprint posted online July 16, 2022:2022.07.13.499978. doi:10.1101/2022.07.13.499978

26. Oken E, Kleinman KP, Rich-Edwards J, Gillman MW. A nearly continuous measure of birth weight for gestational age using a United States national reference. BMC Pediatr. 2003;3:6. doi:10.1186/1471-2431-3-6

27. Reho P, Zhao T, Zhao Y, et al. Maternal epigenetic signatures are associated with small for gestational age births among black women. Epigenetics Insights. 2026;19:e009. doi:10.48130/epi-0026-0006

28. Huang W, Han G, Taylor BD, Neal G, Kochan K, Page RL. Maternal peripheral blood telomere length and preterm birth in African American women: a pilot study. Arch Gynecol Obstet. 2025;311(6):1591–1598. doi:10.1007/s00404-024-07681-1

29. Geronimus AT, Hicken MT, Pearson JA, Seashols SJ, Brown KL, Cruz TD. Do US black women experience stress-related accelerated biological aging? Hum Nat. 2010;21(1):19–38.

30. Ross KM, Carroll JE, Horvath S, Hobel CJ, Coussons-Read ME, Dunkel Schetter C. Epigenetic age and pregnancy outcomes: GrimAge acceleration is associated with shorter gestational length and lower birthweight. Clin Epigenetics. 2020;12(1):120. doi:10.1186/s13148-020-00909-2

